# A Pilot Randomized Trial of Combined Cognitive-Behavioral Therapy and Exercise Training Versus Exercise Training Alone for the Management of Chronic Insomnia in Obstructive Sleep Apnea

**DOI:** 10.1101/2023.03.10.23287115

**Authors:** Amanda Cammalleri, Aurore A. Perrault, Alexandra Hillcoat, Emily Carrese-Chacra, Lukia Tarelli, Rahul Patel, Marc Baltzan, Florian Chouchou, Thien Thanh Dang-Vu, Jean-Philippe Gouin, Veronique Pepin

## Abstract

**Rationale:** Among individuals with obstructive sleep apnea, about 38% will present with comorbid chronic insomnia. Comorbid chronic insomnia and obstructive sleep apnea (COMISA) is associated with poorer daytime functioning and increased morbidity compared to either disorder alone. The treatment of insomnia among individuals with COMISA is suboptimal.

**Objectives:** To assess the effects of an exercise-training intervention alone as well as a combined cognitive behavioral therapy and exercise training on insomnia symptoms in individuals with COMISA.

**Methods:** In a pilot randomized-controlled trial, 19 individuals with COMISA were allocated to one of two arms, each comprising of 2 phases of 8 weeks. The first phase examined the effects of exercise training versus relaxation training on insomnia severity and cardiorespiratory fitness. The second phase compared changes in Insomnia Severity Index (ISI) following exercise training alone (EX) or combined Cognitive Behavioural Therapy for Insomnia and exercise training (CBTi-EX). Self-reported insomnia severity, objective sleep parameters during overnight polysomnography, and cardiopulmonary fitness were collected at baseline, 8 and 16 weeks.

**Results:** Exercise training increased VO_2peak_ and reduced insomnia severity (Hedges’ *g* = 0.82) when compared to relaxation training. Moreover, combined CBTi and exercise training yielded a larger improvement in insomnia severity (Hedges’ *g* =1.48) than exercise training alone (EX). We observed no change in objective sleep measures in both groups.

**Conclusions:** Exercise training leads to moderate improvements in insomnia severity in individuals with COMISA. Combining exercise training with CBTi leads to larger reductions in insomnia symptoms, with effect sizes similar to the effects of CBTi alone in other populations with chronic insomnia.

**Clinical Trial information:** “*Cognitive behavioural therapy combined with exercise training for adults with both insomnia and obstructive sleep apnea*” - (https://doi.org/10.1186/ISRCTN63989489)

## Introduction

Obstructive sleep apnea (OSA) and chronic insomnia are the most common sleep disorders. They each affect about 10% of the general population^1,2^. Both disorders are associated with risk for poor physical and mental health outcomes and are an important societal financial burden^3–5^. OSA is caused by narrowing (hypopnea) or total obstruction (apnea) of the pharyngeal airway during sleep which impairs breathing and airflow. Chronic insomnia involves self-reported difficulty initiating sleep onset, maintaining sleep and/or early awakenings occurring at least 3 nights per week for more than 3 months, and is accompanied by daytime symptoms. Comorbid chronic insomnia and obstructive sleep apnea (COMISA) is a prevalent condition, with about 38% of patients with OSA reporting chronic insomnia ^6–8^. COMISA is associated with worse daytime functioning, lower quality of life, and higher rates of cardiovascular disease and psychiatric issues as well as increased risk of mortality compared to either disorder alone^9–12^.

The treatment of chronic insomnia in COMISA is complex. Although continuous positive airway pressure (CPAP) treatment leads to clinically significant reductions in apnea-hypopnea index and has been shown to improve subjective sleep quality^13^, still about 55% of patients experience persistent insomnia symptoms despite regular CPAP use^14,15^. Furthermore, CPAP acceptance and adherence is often suboptimal; 23%-47% of COMISA patients are not adherent to CPAP in the long-term^16–19^. Notably, those with higher insomnia symptoms typically show lower daily CPAP use 6 months later^20^. Similarly, COMISA patients are less likely to accept and adhere to CPAP therapy compared to those with OSA alone^21–23^, highlighting a need for insomnia-specific interventions among individuals with COMISA.

Cognitive-behavioural therapy for insomnia (CBTi) is considered the first line treatment for chronic insomnia^24^. CBTi is a multimodal sleep-focused psychological intervention aimed at modifying maladaptive thinking and behaviors that contribute to the perpetuation of insomnia^25,26^. CBTi has been shown to decrease self-reported Insomnia Severity Index with medium-to-large effect sizes, even without changes in objective sleep measures, among individuals with chronic insomnia^27–29^ as well as those with COMISA^7,30,31^. While CBTi is the most effective treatment for chronic insomnia to date, 40 to 65% patients with COMISA continue to experience residual or recurring insomnia symptoms after CBTi^32^ or CBTi combined with CPAP^7^. Thus, there is a clear need for novel or adjunct interventions for this uniquely challenging patient population.

Exercise training may be beneficial for COMISA patients, not only for its positive effects on cardiopulmonary fitness but also to improve sleep quality and alleviate insomnia complaints. Aerobic and resistance exercise training interventions have been shown to reduce apnea and hypopnea events and improve oxygen desaturation among patients with OSA^33–38^. Several randomized controlled trials have revealed an increase in self-reported sleep quality following an exercise intervention among patients with OSA^33,34,39^ as well as in individuals with chronic insomnia^40–43^. Specifically, a variety of exercises, including structured moderate-intensity aerobic training, resistance training, tango, and yoga, over periods of 2 to 6 months, led to significant reductions in insomnia severity with moderate effect sizes in individuals suffering from chronic insomnia^42,44–46^. While the effects of regular physical activity on objective sleep measures show varying results, some studies indicate that exercise training may shorten sleep latency and increase sleep efficiency in chronic insomnia^41,42,47^ and OSA^34,36^. In sum, exercise training has been shown to improve sleep quality in OSA and in chronic insomnia alone; however, its effect on insomnia severity has yet to be investigated in the COMISA population.

In addition to improving insomnia symptoms on its own, regular exercise may be a valuable adjunct treatment to combine with CBTi in COMISA. Indeed, the mechanisms of action in exercise training and CBTi are thought to be different. While CBTi targets sleep-related behaviors as well as emotional and cognitive hyperarousal that perpetuates insomnia^48^, exercise is thought to improve objective and subjective sleep by modulating autonomic, metabolic, and inflammatory processes and increasing sleep drive^49^. These complementary mechanisms of action could be mutually beneficial and trigger a virtuous circle, especially in COMISA. There is some evidence that the augmentative effect of exercise training combined with cognitive-behavioural therapy is associated with a larger reduction in anxiety^50^ compared to CBT alone in individuals with anxiety disorders, but the combination of exercise and CBT or CBTi has not been tested in the context of chronic insomnia or COMISA.

The goals of this trial were to examine the impact of exercise training alone and exercise training combined with CBTi on insomnia severity among patients with COMISA. We therefore conducted a pilot randomized-controlled trial with 2 arms consisting of 2 phases of 8 weeks each. In the first phase of the trial, we aimed to examine the effects of 8 weeks of moderate-intensity exercise training (EX group) compared to 8 weeks of self-guided relaxation training (CBTi-EX group) on cardiorespiratory fitness and insomnia severity. Relaxation training was chosen as an active control given that a relaxation intervention is not superior to placebo in relieving insomnia symptoms and improving daytime functioning in insomnia disorder^51–53^. In the second phase, we assessed the effects of 8 weeks of combined CBTi and exercise training (CBTi-EX group) on insomnia severity (ISI) compared to 8 weeks of exercise training alone (EX group). Self-reported insomnia symptoms measured with the Insomnia Severity Index (ISI) questionnaire was the primary outcome. Secondary outcomes included cardiorespiratory fitness (peak oxygen consumption, i.e., VO_2peak_), subjective sleep quality and daytime sleepiness as well as objective assessment of sleep quality using polysomnographic (PSG) recordings (i.e., sleep efficiency, sleep onset latency, wake after sleep onset, sleep fragmentation index, total sleep time). For the first aim, we expected to see a larger reduction in insomnia severity (ISI) and an improvement in cardiopulmonary fitness (greater VO_2peak_) after 8 weeks of exercise training (EX) compared to relaxation training (CBTi-EX). We also expected to see a small effect of the exercise intervention on objective sleep measures (i.e., increased sleep efficiency) compared to relaxation intervention. For the second phase of the study, we hypothesized that the CBTi-EX group would experience larger reductions in insomnia severity after 8 weeks of combined CBTi and exercise training compared to the EX group, which received solely exercise training. We expected improvements in objective sleep (i.e., increased sleep efficiency) in both group 16-weeks post-randomization.

## Material & Methods

### Participants

Participants diagnosed with mild to moderate OSA and complaining of chronic insomnia disorder were included in the study. Participants diagnosed with OSA were recruited through online and print advertisements posted in the community and from physician referrals. They had to have received a diagnosis of mild to moderate OSA by a physician, defined by an apnea-hypopnea index between 5 and 30 per hour. After a preliminary phone screening, a semi-structured in-person interview was conducted by a trained research coordinator to assess the presence of chronic insomnia disorder and other mental disorders using Diagnostic and Statistical Manual of Mental Disorders-5 (DSM-5) criteria. Potentially eligible participants underwent a polysomnography (PSG) recording to habituate to the PSG-setting and lab environment as well as to rule out other sleep disorders (i.e., periodic limb movement >15/h, REM behavioral disorder). Exclusion criteria were as follows: being less than 18 years old, other sleep disorder (i.e., narcolepsy, restless leg syndrome, or REM sleep behavior disorder), medical condition that is a contra-indication for exercise training based on the Physical Activity Readiness Questionnaire^54^, previously received CBTi, major cardiovascular event or intervention (e.g. stroke, myocardial infarct, heart failure, heart surgery, pacemaker, etc.), diabetes, cancer treatment in the last 2 years, epilepsy, neurological disease (e.g., multiple sclerosis, Parkinson’s disease, dementia, etc.), major surgery in the last 3 months, severe mental disorders (e.g., mood disorders and psychotic disorders), alcohol or drug abuse, being pregnant or breastfeeding. Individuals taking hypnotic medications were asked to stop using them for at least two weeks before each assessment and were encouraged not to use them during the intervention. While the use of a CPAP machine was encouraged, it was not mandatory during study assessments. Participants using CPAP had to have used CPAP for at least three months before being randomized for the study. Lastly, individuals who reported participating in over 150 minutes per week of moderate-to-vigorous exercise were excluded to avoid potential ceiling effects in gains from exercise training. All participants signed a written informed consent form before entering the study, which was approved by the Concordia University Human Research Ethics Committee.

### Protocol

Within a month of the initial PSG screening/adaptation night, eligible participants completed baseline, pre-treatment assessments (T1), which included a second PSG, a cardiopulmonary exercise test (CPET) in-lab, and self-report questionnaires. Following these baseline assessments, participants were randomized to either 16 weeks exercise training (EX group) or 8 weeks of self-guided relaxation followed by 8 weeks of combined CBTi and exercise training (CBTi-EX group). The randomization allocation ratio was 1:1 with 4 participants per block (http://www.jerrydallal.com/random) to preserve the blind random assignment while minimizing imbalance between groups in a pilot study. To maintain allocation concealment, results were contained in sealed envelopes prepared prior to study initiation and opened sequentially in the presence of both the patient and research assistant. At 8 weeks post-randomization (T2) and 16 weeks post-randomization (T3), participants from both groups underwent the same assessment protocol as they did at baseline (i.e., PSG, CPET, questionnaires – **Figure.1**). The present project was registered as a clinical trial (ISRCTN63989489 - https://doi.org/10.1186/ISRCTN63989489).

### Interventions

#### Exercise training

Exercise training involved 60-minute sessions of structured moderate-intensity aerobic exercise combined with individualized resistance training, three times per week. The intervention program was the same for both groups but lasted 16 weeks for the EX group and 8 weeks for the CBTi-EX (during the second intervention phase). Aerobic training consisted of 5 minutes of warm-up, followed by 30 minutes of aerobic exercise performed at the heart rate (HR) associated with the ventilatory threshold as determined from the prior CPET, and finally 5 minutes of cool-down. Resistance training consisted of one set of 12-15 repetitions for 6-8 different exercises. Of the three weekly training sessions, one was conducted under the direct supervision of a trained exercise physiologist and two were unsupervised, taking place at the participant’s home or community. Due to the COVID-19 pandemic, 3 participants received their direct supervision of exercise training via videoconferencing.

#### Relaxation training

Relaxation training involved self-guided sessions of digital audio recordings at least 3 times per week during 8 weeks for the CBTi-EX group only (immediately post-randomization). Relaxation audio recordings consisted of diaphragmatic breathing exercises, progressive muscle relaxation, and guided imagery^52,55,56^.

#### Cognitive-Behavioral Therapy for insomnia (CBTi)

During the second intervention phase (from T2 to T3), participants in the CBTi-EX group underwent 6 sessions of CBTi with a trained psychologist over 8 weeks (4 weekly sessions followed by 1 session every 2-3 weeks). The CBTi program consisted of psychoeducation about sleep and circadian rhythms, stimulus control, sleep restriction, sleep hygiene, cognitive therapy, and relaxation based on Morin and Espie^57^. Due to the COVID-19 pandemic, 2 participants underwent CBTi therapy via videoconferencing.

### Measures

#### Self-Reported Questionnaires

##### Insomnia Severity Index (ISI)

The ISI is a widely used 7-item self-report questionnaire examining the nature, severity, and impact of current insomnia symptoms^58,59^. The total score ranges from 0 to 28, with higher scores indicating more severe insomnia. Its Cronbach’s α is .0.68 in the current study. Participants were considered responders when the total ISI score decreased by over 8 points from one assessment to the next. Participants were considered in remission when the total ISI score was below 8.

##### Pittsburgh Sleep Quality Index (PSQI)

The PSQI is a self-reported measure of general sleep quality^60^, with a total score from 0 to 21, with higher scores indicating worse sleep quality. The Cronbach’s α in the current sample is 0.83.

##### Epworth Sleepiness Scale (ESS)

The ESS is a self-reported questionnaire to measure daytime fatigue and sleepiness. It includes 8 items scored from 0 to 3 indicating chances of dozing off or falling asleep while engaged in eight different activities. Total score ranges from 0 to 24 with higher scores indicating more severe daytime sleepiness. The Cronbach’s α in the current sample is 0.88.

#### Cardiopulmonary Exercise Test (CPET)

CPETs were conducted to rule out the presence of any cardiorespiratory condition that would interfere with exercise participation (exclusion criterion), to determine the target intensity for exercise training, and to assess the physiological impact of the exercise intervention. The exercise test was a symptom-limited incremental test conducted on a cycle ergometer (Corvial, Lode, Groningen, The Netherlands) following the Jones protocol^61^. It was administered by trained and certified clinical exercise physiologists. Measurements included resting, exercising, and recovering hemodynamic response (heart rate via electrocardiography and blood pressure via automated BP monitoring), gas exchange and ventilatory response (oxygen consumption, carbon dioxide excretion, minute ventilation, breathing pattern, inspiratory capacity via Medgraphics CardiO2 Metabolic Cart, MCG Diagnostics, Saint Paul, MN, USA), oxygen saturation via pulse oximetry, and ratings of perceived exertion and dyspnea^62^. Exercise capacity was defined as the highest workload (in Watts) achieved and maintained for at least 30 seconds. Peak oxygen consumption (VO_2peak_) was averaged from the last 30 seconds of the testing phase. The workload, oxygen consumption, and heart rate at which the ventilatory threshold occurred were identified using the V-slope approach. Height and weight were taken by a certified nurse, and body mass index (BMI) was calculated from this information accordingly (kg/m^2^).

#### Polysomnographic (PSG) recording

Participants underwent four whole-night PSG recording (screening night, T1, T2, T3). The EOG, EMG and 14 EEG scalp electrodes (Fpz, F3, Fz, F4, C3, Cz, C4, P3, Pz, P4, O1, O2, M1, M2) were placed according to the international 10-20 system. Screening PSG also included leg EMG electrodes to screen for periodic limb movements (PLMS index > 15/hr), as an exclusion criterion. The PSG signal was recorded with a Somnomedics amplifier (Somnomedics GmbH, Germany), sampled at 512 Hz. EEG recordings were referenced online to Pz, and for the offline analyses the EEG signals were re-referenced to the contra-lateral mastoids (M1, M2). All sleep scoring and analyses were conducted using the Wonambi python toolbox (https://wonambi-python.github.io). For each recording, two scorers blind to allocation determined the different sleep stages (NREM 1, 2, 3, REM sleep and wake) according to the AASM rules for each recorded night of sleep^63^. Sleep onset latency (SOL; min), wake after sleep onset (WASO; min), total sleep time (TST; min), sleep fragmentation index (SFI; % : number of stage shifts to lighter stage/ TST x100), sleep efficiency (SE; % : TST/ time spent in bed x100), were extracted from the sleep scoring. Using pulse oximeter, we monitored and extracted oxygen saturation (SpO_2_) as well as percentage of time spent in SpO_2_ below 90% during total sleep time.

Note that the COVID-19 pandemic began while this study was in progress and impacted in-person data collection, specifically for CPETs and PSGs, for several months (see **Figure 1B** for attrition of each measure).

**Figure 1.**
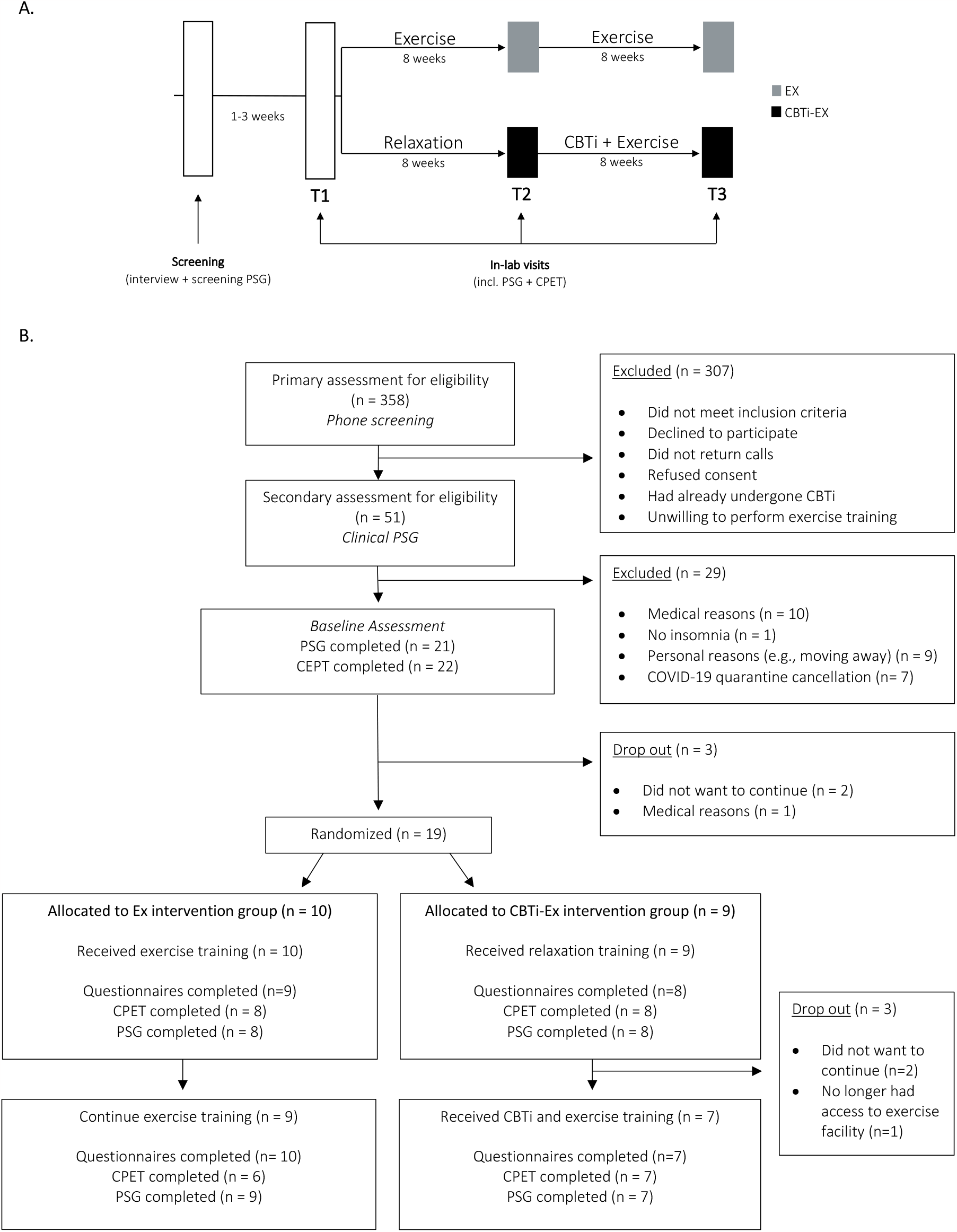
Study design and participant flowchart. (A) At the pre-treatment assessment (T1) participants underwent a baseline assessment (PSG recording, CPET and self-reported questionnaires) then were randomized into either a 2×8 weeks of exercise training alone (EX group - grey) or an 8-week relaxation training followed by 8 weeks of combined CBTi and exercise training (CBTi-Ex group - black). After 8 weeks (T2) and 16 weeks post-randomization (T3), participants underwent the same assessments as baseline. (B) Consort flow chart of participants PSG, polysomnography; AHI, apnea-hypopnea index; PLM, periodic limb movements during sleep; CEPT, cardiopulmonary exercise test; CBTi, Cognitive-Behavioral Therapy for insomnia.

### Statistical analyses

Statistical analyses were performed using RStudio 1.2.50 (RStudio, Inc., Boston, MA) and R packages (e.g., car, emmeans, sjstats, forcats, effsize). An Intent-to-Treat analysis was conducted on our primary outcome measure (i.e., ISI score) using repeated measures hierarchical linear modeling (HLM) analysis to evaluate changes in insomnia severity. The statistical model included Group, Time and the interaction term Group by Time. HLM included random effects for participants and residual error. Due to the COVID-19 pandemic, we were not able to collect complete data for each subject for certain measures. We decided to conduct per protocol analyses to investigate the change of our secondary outcomes (i.e., cardiorespiratory fitness) and exploratory measures (i.e., PSQI, ESS, PSG-determined sleep variables) across time (within subjects) and between groups. To that end, we performed mixed-model analysis of variance (ANOVA) with Group as a between-subject factor (EX group vs CBTi-EX group), Time as a within-subject factor (T1 vs T2 vs T3) and assessed Group by Time interaction. Post-hoc tests of main and interaction effects were also performed when necessary, using Bonferonni correction. Level of significance was set at p < 0.05 and degrees of freedom were corrected according to the Greenhouse–Geisser method. Due to the small sample size, we reported within-effect sizes using Hedges’s *g* (corrects for small sample size). Visual inspection and the Shapiro test were used to determine normality of the data and homogeneity of variance was tested with Levene tests. Non-parametrical version of the analyses (e.g., Wald-Type Statistic) were used when the assumptions of the linear model were not met.

## Results

Nineteen individuals with confirmed COMISA were randomized into either 16 weeks of structured exercise training (EX group; N=10) or 8 weeks of self-guided relaxation training followed by 8 weeks of combined CBTi and structured exercise training (CBTi-EX group; N=9). Participants were, on average, middle-aged (56 ± 13 years old), predominantly female (59%), with BMI values predominantly in the overweight to obese range (M(SD) = 31.0 ± 8.3). The majority (68%) used a CPAP throughout the study. There were no significant group differences in age, BMI, ISI, VO_2peak_, or CPAP use at baseline (all *p*>.05; see **Table 1** for demographics).

**Table 1.** Demographics

|  |  | **All Participants**<br>**(N=19)** | **EX Group**<br>**(N=10)** | **CBTi-EX Group**<br>**(N=9)** |
| --- | --- | --- | --- | --- |
| **Age** | Mean ± sd | $56 \pm 13$ | $61 \pm 10$ | $50 \pm 14$ |
|  | Median | 57 | 64 | 55 |
|  | Range | 28-70 | 36-70 | 28-67 |
| **BMI** | Mean ± sd | $31.0 \pm 8.3$ | $28.3 \pm 6.8$ | $34.1 \pm 9.5$ |
|  | Median | 28.4 | 27.0 | 30.5 |
|  | Range | 17.9-54.2 | 17.9-40.5 | 24.3-54.2 |
| **ISI score** | Mean ± sd | $16.6 \pm 4.7$ | $16.5 \pm 5.4$ | $16.8 \pm 4.2$ |
|  | Median | 17 | 17 | 17 |
|  | Range | 8-25 | 8-25 | 10-25 |
| **$VO_2$ peak**<br>**(ml/kg/min)** | Mean ± sd | $20.7 \pm 7.6$ | $19.58 \pm 6.41$ | $21.97 \pm 8.83$ |
|  | Median | 19.3 | 19.5 | 19.3 |
|  | Range | 11.0-43.2 | 11.0-28.7 | 13.5-43.2 |
| **Female** | n (%) | 11 (57.8) | 6 (60) | 5 (55.5) |
| **CPAP User** | n | 13 | 8 | 5 |

*BMI, body mass index; CPAP, continuous positive airway pressure machine, ISI, insomnia severity index*

### Phase 1: exercise training vs. relaxation training

At the end of the first intervention phase, 8 weeks post-randomization (T2), there was a Group by Time interaction on VO_2peak_ change (*F*(1,14)=10.1, *p*=.007), whereby an increase in VO_2peak_ was detected in the EX group (N=8) after 8 weeks of structured exercise training (*p*=.03, *g’*=-0.41) while no such change was observed in the CBTi-EX group (N=8) undergoing relaxation training (*p*=.07, *g’*<0.1; **Table S1**). These findings attest that our exercise intervention was effective at improving cardiorespiratory fitness.

Regarding changes in insomnia severity, we found a main effect of Time (*F*(1,16)=10.4, *p*=.005) and a trend for a Group by Time interaction (*F*(1,16)=3.9, *p*=.07) due to the large reduction in ISI score at T2 in the EX group only. On average, the EX group reduced their ISI score by 4.7 ± 3.2 points after 8 weeks of exercise training (*p*=.001, *g’*=0.82) while the CBTi-EX group who completed 8 weeks of self-guided relaxation did not show a significant change in ISI score (−1.1 ± 4.4 points; *p*=.49, *g’*=0.16) (**Figure 2**). While the EX group displayed larger reduction in ISI than the CBTi-EX group at T2, none of them were considered responders (i.e., ISI decreasing by ≥ 8 points) 8 weeks post-randomization. However, one participant (12.5%) in the EX group was considered in remission (i.e., ISI score <8) at T2 (**Figure S1**).

**Figure 2.**
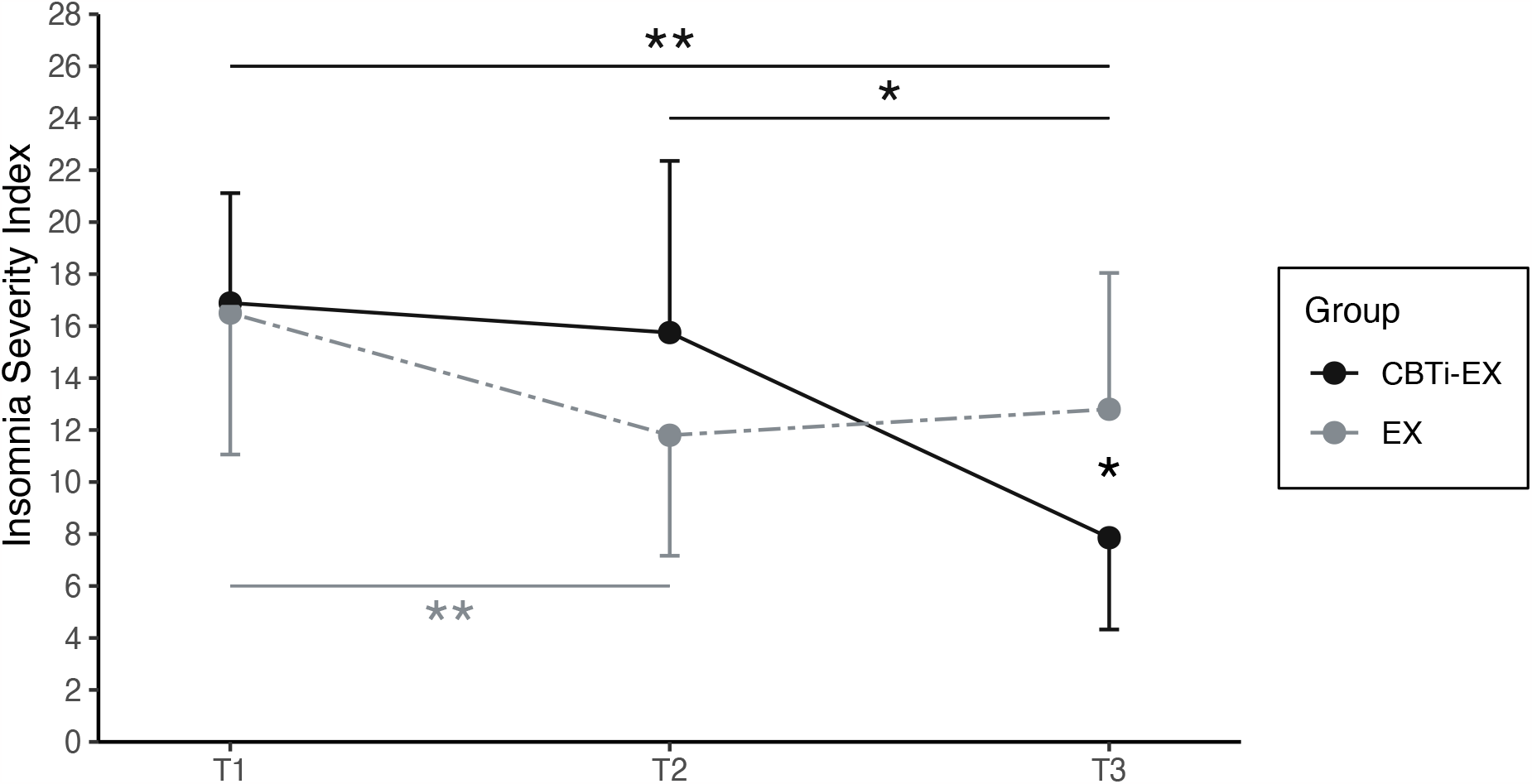
Change in insomnia severity. Mean± SD ISI score at baseline (T1), 8 weeks post-randomization (T2) and 16 weeks post-randomization (T3) for the EX group (grey; N=10) and CBTi-EX group (black; N=7) Asterisks represent significance (p): *<.05 **<.001

### Phase 2 : exercise training vs. combined CBTi and exercise training

Due to COVID-19 quarantine restrictions, we were unable to conduct CPETs during several months. As a result, T2 and/or T3 VO_2peak_ measurements were missing for 7 (37%) participants. We were therefore not powered to statistically analyze changes in VO_2peak_ from T2-T3 and thus cannot confirm the effect of the exercise intervention on fitness in the CBTi-EX group versus the EX group (**Table S1**).

HLM analyses on the trajectory of ISI score at 16 weeks post-randomization (T3; end of the second intervention phase - EX N=10; CBTi-Ex N=7) revealed a significant Group by Time interaction (chi-square *p*<.001) and main effect of Time (chi-square *p*<.001; **Figure 2**). While the CBTi-EX group reported no change in insomnia severity after the 8 weeks relaxation program, they showed significant reductions in ISI score after 8 weeks of combined CBTi and exercise training (T2 vs T3; -8.9 ± 7.7pt; *b(SE)* = -9.7, (1.7), *t*=-5.54, *p*=.023, *g’*=1.48). Meanwhile, in the EX group, following the reduction in ISI from T1 to T2, there was no further change in insomnia severity from T2 to T3 (1± 3.9pt; *beta* = 6.01, *SE* = 2.31, *t*=2.59, *p*=.88, *g’*<0.1).

On average, from baseline (T1) to the end of the second intervention phase (T3; 16 weeks post-randomization), the CBTi-EX group reduced their ISI by 10 ± 3.7 points (*p*<.001, *g’*=2.34) while the EX group reduced ISI score by 3.7 points ± 5.4 (*p*=.058, *g’*=0.63). At end of the second intervention phase, 6 out of 7 participants (85.7%) who completed the CBTi-EX intervention were considered as responders and 3 (43%) were considered in remission. Meanwhile, in the EX group, 2 participants out of 10 (20%) were considered as responders as well as in remission at T3 (**Figure S1**).

### Exploratory analyses of change in sleep quality measured with objective and subjective assessment

At 16 weeks post-randomization (T3; CBTi-Ex N=7; EX N=5), per protocol investigation of sleep architecture using PSG recording (e.g., sleep efficiency, total sleep time, sleep latency, time spent awake after sleep onset, sleep fragmentation index) did not reveal any main effects or interaction Group by Time (all *p* >.05) as well as small-to-no effect sizes from baseline (T1) to 8 weeks (T2) or 16 weeks (T3) post-randomization (all *g’*<0.1). There was also no change in mean nocturnal oxygen desaturation (SpO_2_) and time spent with SpO_2_ below 90% in both groups at T2 and T3 (all *p*>.05; **Table S2)**.

Per protocol assessment of change in sleep quality using the PSQI (CBTi-EX N=6; EX N=7) revealed significant effect of Time (*F*(2,22)= 7.9; *p*=.003), but no effect of Group (*F*(1,11)= 2.8; *p*=.11) or Group by Time interaction (*F*(2,22)= 0.13; *p*=.87). The EX group did not significantly reduce PSQI score from T1 to T2 (*p*=.85, *g’*<0.1) or from T2 to T3 (*p*=.1, *g’*=0.72). However, overall, after 16 weeks of exercise training, they displayed a large reduction in PSQI score compared to baseline (T1 vs T3, p=.003, g’=0.89). Furthermore, although there was no change in PSQI scores after 8 weeks of relaxation training (T1 vs T2; *p*=.75, *g’*=-0.12), there was a large reduction in PSQI after 8 weeks of combined CBTi and exercise training in the CBTi-EX group (T2 vs T3; *p*=.04, *g’*=1.2; **Table S3**).

Finally, there was no significant Group by Time interaction or main effects of Time or Group (all *p*>.05) on daytime sleepiness using the ESS questionnaire (CBTi-EX N=6; EX N=8). There was a medium effect size (*g’*=0.59) concerning change in ESS after 8 weeks of exercise training (T1 vs T2; -2.8 ± 2.2 points) in the EX group but no change from T2 to T3 (*g’*=-0.1; T1 vs T3: *g’*=0.32; **Table S3)**. Participants in the CBTi-EX group had no change in daytime sleepiness from T1 to T2 (*g’*<0.1) but experienced a small reduction in daytime sleepiness from T2 to T3, (−1.66 ± 3.55 points; *g’*=0.23; T1 vs T3: *g’*=0.22; **Table S3**).

## Discussion

Chronic insomnia complaints are reported by more than 38% of people with OSA, and this comorbid condition (COMISA) is associated with worse daytime functioning and morbidity profile compared to either disorder alone. Here, we reported the effects of exercise training and exercise training combined with CBTi on insomnia severity among COMISA patients. In this pilot randomized controlled trial, a structured exercise-training intervention led to increases in cardiorespiratory fitness and reductions in insomnia severity compared to relaxation training in people with confirmed COMISA. We also found that combined CBTi and exercise training (CBTi-EX) yields larger reductions in insomnia complaints compared to exercise alone (EX). Finally, we found no change in objective sleep measures assessed with polysomnographic recordings in either group.

After 8 weeks of exercise training, individuals with COMISA who exercised (EX group) demonstrated a moderate improvement in their cardiorespiratory fitness (indicated by a significant increase in VO_2peak_ during CPET) whereas those who did 8 weeks of relaxation training (CBTi-EX group) did not display such a change. While improvements in cardiorespiratory fitness after exercise training have been reported in OSA alone^64,65^, our results extend these findings to individuals with COMISA. Furthermore, the fact that our exercise intervention was of moderate intensity and followed a hybrid format of supervised and unsupervised sessions suggests that it would be applicable in a ‘real world’ setting.

The EX group reported a moderate reduction in ISI score (*g’*=0.82 at T2 and *g’*=0.63) as well as a small change in daytime sleepiness (*g’*=0.59 at T2 and *g’*=0.32 at T3) after 8 weeks and 16 weeks of exercise training. Such small-to-moderate change in insomnia severity is consistent with other studies examining the effects of regular exercise among individuals with chronic insomnia^42,44–46^. In the COMISA population, implementation of regular moderate physical activity could be beneficial not only for cardiovascular health but also to reduce subjective sleep complaints. However, it is important to note that we did not find any changes in objective sleep measures (i.e., sleep duration, efficiency, latency, fragmentation). While some studies have reported shorter sleep onset latency and improved sleep efficiency in chronic insomnia^41,42,47^ and OSA^34,36^ after exercise training interventions, the effects of regular exercise training on objective sleep measures remain debated as many studies have reported small-to-no effects^40,45,66,67^ and suggests a complex interaction between time of day as well as type and intensity of exercise training to determine the best exercise prescription to improve sleep quality^68^.

Consistent with the extensive literature on the efficacy of CBTi in chronic insomnia^28^, we reported a large improvement (*g’*=1.48) in insomnia severity (ISI) after 8 weeks of combined CBTi with exercise training (CBTi-EX group) compared to 16 weeks of exercise alone (*g’*= 0.89; EX group). Although there is substantially less research in the COMISA population, our results are comparable to what has been found recently by Sweetman and colleagues^7,31^, who reported a large reduction in ISI score after a 4-week CBTi program compared to treatment-as-usual among COMISA patients. Furthermore, the beneficial response rate (86%) and remission rate (43%) are similar to those found in previous trials in chronic insomnia^28^ and COMISA^7,30,31^. Despite changes in subjective insomnia severity, we found no change in objective sleep (i.e., assessed by PSG recording) in both groups. Thus, combined CBTi and exercise training appears to primarily improve self-reported measures of sleep satisfaction, and subjective insomnia symptoms rather than objective measures of sleep quality. Such results are in line with those reported in studies investigating the effects of CBTi alone on objective and subjective sleep measures in chronic insomnia alone^27,28,69^.

The effect size of the combined CBTi and exercise training intervention on insomnia severity is similar to the effect sizes of CBTi alone observed in other studies^7,30,31^. Although our study design did not allow us to disentangle the effects of CBTi alone, the effect sizes obtained from this small pilot trial do not suggest that the combination of CBTi with exercise training leads to larger reduction in insomnia compared to CBTi alone among COMISA patients. Nonetheless, our results indicate that a combined CBTi and exercise training intervention is feasible and acceptable for some COMISA patients and likely leads to improvements in both cardiopulmonary fitness and insomnia severity. However, it is important to acknowledge the possible sampling biases associated with our stringent inclusion and exclusion criteria as well as participants’ motivation to voluntarily enroll in an exercise training intervention (3 times a week).

Furthermore, although CBTi combined with exercise training was associated with a larger reduction in insomnia severity than 16 weeks of exercise alone, it is important to note that both groups displayed moderate-to-large changes in insomnia severity 16 weeks post-randomization, suggesting that both interventions can be effectively implemented for the management of insomnia complaints for COMISA patients. Despite the strongly documented efficacy of CBTi, there is limited access to CBTi therapy as it requires considerable human (i.e., availability of trained professionals), time (i.e., 2h-sessions during 6 to 10 weeks) and financial resources, which constitute important barriers to its clinical widespread use^70–72^. Our results suggest that the implementation of regular moderate exercise training could be approached as an initial and/or complementary solution with moderate effect in the management of chronic insomnia in COMISA, but similar studies with greater sample sizes are warranted.

Some study limitations affect the interpretation and generalizability of the findings. First, our sample size is small. The COVID-19 pandemic impacted recruitment, in-person data collection, and intervention delivery. However, we were able to collect self-reported questionnaires and ensure participation in weekly CBTi sessions and exercise training via videoconference. Additionally, since the CPAP use did not affect eligibility for study inclusion, the number of CPAP users differs across treatment arms. Relatedly, more than half of the participants used CPAP (including during their nights at the lab) and, consequently, we were not able to investigate the effects of exercise training on the number of apnea and hypopnea events. Such effects should be investigated in future studies on the impact of regular exercise in COMISA, as regular exercise training has been shown to improve breathing difficulties in OSA alone^33–37,66,73^. Finally, despite the small sample size, the use of a randomized controlled study with 2-arms that includes two phases of intervention each allows for the examination of within-individual effects, thus accounting for many inter-individual differences in sleep and exercise.

This pilot randomized controlled study showed that moderate-intensity exercise training improved cardiorespiratory fitness in individuals with COMISA. It also showed promising effects of regular exercise training alone (moderate) and combined with a CBTi program (large) on insomnia severity in COMISA. None of the interventions (exercise alone or combined CBTi and exercise training) showed improvements in objective sleep measures when assessed with gold-standard PSG recordings. Future studies should examine whether a sequential or combined delivery of exercise training and CBTi is optimal to improve sleep and cardiopulmonary outcomes among individuals with COMISA.

## Supporting information

Supplemental Results

## Data Availability

All data produced in the present study are available upon reasonable request to the authors

## Author contribution statement

Conceptualization: VP, TDV, JPG; Data collection: AC, AAP, LT, ECC, AH, FC; Data curation: AC, AAP, AH, ECC, VP; Formal analysis: AC, AAP; Methodology: AAP, AC, VP, JPG; Funding acquisition: VP, TDV, JPG, FC; Project coordination/administration: ECC, LT, AH, AC, AAP, VP, JPG, TDV; Visualization: AAP, AC; Writing: AC, AAP, VP, JPG, TDV.

## Acknowledgments

This research was supported by grants from the (Canadian) Lung Association Program: Breathing as One Allied Health Research Grant (grant number not provided), the PERFORM Centre, Concordia University Program: Innovative Research Project in Preventive Health (grant number not provided) and the Canadian Research Continuity Emergency Fund (SCV108) to VP; and by the Canada Research Chair of JPG.

We acknowledge the contributions of the following students who assisted in participants’ recruitment, data collection and data preprocessing: Kajamathy Subramaniam, Emma-Maria Phillips, Rachel Hu and all the volunteers. We acknowledge the contribution of the psychologists (Rosemarie Perrault, Caroline Desrosiers) who provided CBTi and the clinician (Chantal Lafond) who referred patients. We also thank our sleep technologists Madeline Dickson and Elinah Mozhentiy, and Liza Perez from the Clinique SomnoMed for their contribution to the setup of sleep recordings. Finally, we would like to thank the participants for giving their time and energy into this research study.

