## Supplemental Results for "A Pilot Randomized Trial of Combined Cognitive-Behavioral Therapy and Exercise Training Versus Exercise Training Alone for the Management of Chronic Insomnia in Obstructive Sleep Apnea"

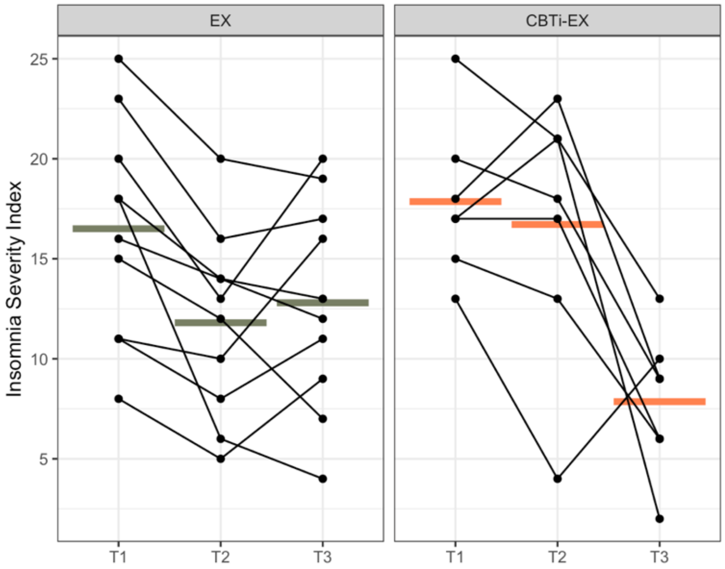


**Figure S1 – Change in insomnia severity**

*Individual changes (black line) and mean (green and orange bars) ISI score at baseline (T1), 8-weeks post-randomization (T2) and 16-weeks post-randomization (T3) for the EX group (left panel; N=8) and CBTi-EX group (right panel; N=6)*

**Table S1 – Change in cardiorespiratory fitness**

**EX Group**

| Participant Number | T1 | T2 | T3 |
|---|---|---|---|
| E1 | 20.4 | 21.8 | NA[1] |
| E2 | 23.3 | 29.5 | 25.3 |
| E3 | 14.6 | 18.8 | NA[1] |
| E4 | 13.2 | 20.9 | 20.1 |
| E5 | 25.5 | 27.3 | 28.4 |
| E6 | 28.7 | 26.6 | 28 |
| E7 | 27.2 | 29.7 | 34.6 |
| E8 | 13.3 | 14.3 | NA[3] |
| E9 | 11 | NA[2] | NA[2] |
| E10 | 18.6 | NA[2] | 24.7 |
| *Mean* | *19.58* | *23.6125* | *26.85* |
| *SD* | *6.41* | *5.54* | *4.82* |

**CBTi-EX Group**

| Participant Number | T1 | T2 | T3 |
|---|---|---|---|
| C11 | 21.6 | 20.8 | NA[4] |
| C12 | 43.2 | 40.5 | 44 |
| C13 | 19 | 16.4 | 16.9 |
| C14 | 14.7 | 14.8 | 17 |
| C15 | 17.6 | 16.3 | 14.8 |
| C16 | 19.3 | 17.6 | 19.7 |
| C17 | 13.5 | 14.6 | 16.9 |
| C18 | 25.2 | 25.2 | NA[5] |
| C19 | 23.6 | 32.2[6] | 27.4 |
| *Mean* | *21.97* | *22.04* | *22.39* |
| *SD* | *8.83* | *9* | *10.38* |

*Individual changes and mean (± SD) for $VO_2$ peak (ml/kg/min) at baseline (T1), 8-weeks post-randomization (T2) and 16-weeks post-randomization (T3) for the EX group (left panel) and CBTi-EX group (right panel)*

*NA[1]- Could not come in for CPET for medical reasons*

*NA[2]- Could not come in for CPET due to COVID quarantine*

*NA[3]- Could not come in for CPET due to personal reasons*

*NA[4]- Did not exercise from T2- T3 due to lack of accessibility. Was not included in any analyses.*

*NA[5]- Could not come in for CPET due to COVID quarantine*

*[6]-CPET conducted outside the time window prescribed by the protocol (9 months instead of 8-weeks) due to COVID restrictions*

**Table S2 – Change in objective sleep (PSG-related measures).**

| PSG measures | T1 | T2 | T3 | Group*Time effect | Group effect | Time effect |
|---|---|---|---|---|---|---|
| *Sleep Efficiency (%)* | | | | | | |
| EX | 75.5 ± 11.5 | 76.1 ± 11.8 | 68.0 ± 5.2 | S(2,26) =2.8 | S(1,13) =3.7 | S(2,26) =3.2 |
| CBTi-EX | 82.2 ± 14.8 | 78.4 ± 11.8 | 80.7 ± 11.1 | p=.24 | p=.054 | p=.19 |
| *Total Sleep Time (min)* | | | | | | |
| EX | 357.1 ± 73.6 | 343.0 ± 50.2 | 316.3 ± 46.6 | F(2,26) =0.52 | F(1,13) =0.65 | F(2,26) =0.67 |
| CBTi-EX | 365.0 ± 76.5 | 344.6 ± 62.7 | 358.1 ± 61.4 | p=.6 | p=.43 | p=.51 |
| *Sleep Onset Latency (min)* | | | | | | |
| EX | 13.6 ± 15.9 | 19.0 ± 23.4 | 16.9 ± 12.4 | S(2,26) =0.75 | S(1,13) =1.21 | S(2,26) =1.89 |
| CBTi-EX | 14.0 ± 6.3 | 24.3 ± 19.5 | 27.3 ± 34.5 | p=.68 | p=.27 | p=.38 |
| *Wake After Sleep Onset (min)* | | | | | | |
| EX | 86.6 ± 54.9 | 84.7 ± 53.3 | 108.7 ± 62.3 | F(2,26) =0.34 | F(1,13) =4.8 | F(2,26) =0.36 |
| CBTi-EX | 61.8 ± 59.2 | 51.9 ± 29.4 | 56.5 ± 17.6 | p=.71 | p=.05 | p=.7 |
| *Sleep Fragmentation Index (SFI)* | | | | | | |
| EX | 12.3 ± 6.9 | 13.4 ± 5.5 | 12.8 ± 6.8 | F(2,26) =1.2 | F(1,23) =0.3 | F(2,26) =0.6 |
| CBTi-EX | 11.1 ± 2.8 | 10.4 ± 3.0 | 12.3 ± 3.6 | p=.3 | p=.55 | p=.54 |
| *Mean SpO2* | | | | | | |
| EX | 90.4 ± 14.8 | 95.3 ± 1.1 | 94.2 ± 3.2 | F(2,26) =0.38 | F(1,13) =0.37 | F(2,26) =0.51 |
| CBTi-EX | 95 ± 1.1 | 95.4 ± 1.2 | 94.6 ± 0.6 | p=.56 | p=.55 | p=.5 |
| *SpO2 <90 (%)* | | | | | | |
| EX | 5.7 ± 15.3 | 0.52 ± 0.68 | 11.3 ± 29.9 | F(2,26) =0.27 | F(1,13) =0.24 | F(2,26) =0.44 |
| CBTi-EX | 1.99 ± 3.9 | 2.2 ± 4.4 | 4.34 ± 4.7 | p=.66 | p=.63 | p=.55 |

*Mean (±SD) PSG-extracted measures at baseline (T1), 8-weeks post-randomization (T2) and 16-weeks post-randomization (T3); CBTi-EX N=7, EX N=8*

**Table S3 – Change in self-reported sleep quality and sleepiness.**

| Questionnaires | T1 | T2 | T3 | Group*Time effect | Group effect | Time effect |
|---|---|---|---|---|---|---|
| *PSQI* | | | | | | |
| EX | 10.8 ± 3.3 | 10.5 ± 3.9 | **7.7 ± 2.7*** | F(2,22) =0.13 | F(1,11) =2.8 | **F(2,22) =0.13** |
| CBTi-EX | 8.5 ± 3.1 | 8.8 ± 3.3 | **5 ± 1.6*** | p=.87 | p=.11 | **p=.003*** |
| *ESS* | | | | | | |
| EX | 10.5 ± 4.6 | 7.75 ± 4.3 | 8.5 ± 5.8 | F(2,24) =1.36 | F(1,12) =0.55 | F(2,24) =1.84 |
| CBTi-EX | 11.33 ± 5 | 11.5 ± 5.9 | 9.83 ± 6.2 | p=.27 | p=.47 | p=.18 |

*Mean (±SD) scores at baseline (T1), 8-weeks post-randomization (T2) and 16-weeks post-randomization (T3); PSQI CBTi-EX N=6, EX N=7; ESS CBTi-EX N=6, EX N=8*
*PSQI, Pittsburgh Sleep Quality Index; ESS, Epworth Sleepiness Scale*
